# AD-NET: Age-adjust neural network for improved MCI to AD conversion prediction

**DOI:** 10.1101/2020.04.24.20074195

**Authors:** Fei Gao, Hyunsoo Yoon, Yanzhe Xu, Dhruman Goradia, Ji Luo, Teresa Wu, Yi Su, for the Alzheimer’s Disease Neuroimaging Initiative

## Abstract

The prediction of Mild Cognitive Impairment (MCI) patients who are at higher risk converting to Alzheimer’s Disease (AD) is critical for effective intervention and patient selection in clinical trials. Different biomarkers including neuroimaging have been developed to serve the purpose. With extensive methodology development efforts on neuroimaging, an emerging field is deep learning research. One great challenge facing deep learning is the limited medical imaging data available. To address the issue, researchers explore the use of transfer learning to extend the applicability of deep models on neuroimaging research for AD diagnosis and prognosis. Existing transfer learning models mostly focus on transferring the features from the pre-training into the fine-tuning stage. Recognizing the advantages of the knowledge gained during the pre-training, we propose an AD-NET (Age-adjust neural network) with the pre-training model serving two purposes: extracting and transferring features; and obtaining and transferring knowledge. Specifically, the knowledge being transferred in this research is an age-related surrogate biomarker. To evaluate the effectiveness of the proposed approach, AD-NET is compared with 8 classification models from literature using the same public neuroimaging dataset. Experimental results show that the proposed AD-NET outperforms the competing models in predicting the MCI patients at risk for conversion to the AD stage.

## 1 Introduction

Alzheimer’s disease (AD) is one of the most common progressive neurodegenerative diseases in elderly patients. Over 5.5 million Americans presently suffer from AD, and the number is expected to increase to 16 million by 2050 with projected healthcare costs reaching $1.2 trillion (Alzheimer’s Association 2016). AD is characterized by a long preclinical stage with the slow progression of AD related pathologies without clinical symptoms, while substantial neuronal loss has already happened when cognitive changes can be detected (Long, J. M., & Holtzman, D. M., 2019). Early detection is critical for AD because it is commonly believed that this is when the intervention can be more effective before irreversible brain damage occurs (Frost, S. et al., 2013). Thus, Mild Cognitive Impairment (MCI), a pre-dementia stage, has been of great interest in both AD research and clinical practices. MCI is the stage when the individual has greater cognitive decline than expected from normal aging but has not shown noticeable interruptions from the daily activities (Selkoe, D. J. (1997). Studies show that MCI patients with memory complaints and deficits (amnestic mild cognitive impairment) have a higher risk of progression to AD (Gauthier, S. et al., 2006). The ability to identify patients who will have faster cognitive decline and are at a higher risk of converting to clinical AD will facilitate the development of effective treatments by optimizing cohort selection, and allow better management of the disease (Aisen, P.S. et al., 2011). This is a non-trivial task. Fortunately, recent studies have demonstrated that neuroimages can more sensitively and consistently measure disease progression than cognitive assessment (F. Li et al., 2018). Imaging biomarkers as the objective and quantitative criteria have been intensively studied as potential means for AD early detection.

Most research on AD imaging biomarkers focuses on discovering the features directly measured from the images such as structural Magnetic Resonance Imaging (MRI), Positron Emission Tomography (PET), and resting-state functional MRI (fMRI) (Fox, N. C. et al., 2000; Chupin, M., et al. 2009; Eskildsen, S. F., 2013; Filippi, M. et al., 2012; Pike, K. E. et al., 2007; Li, H. J. et al., 2015; Yamada, T. et al., 2017). Biomarker discovery requires joint efforts from predictive modeling and medicine domain knowledge. Earlier works on modeling have been mainly related to machine learning pipeline, where feature extraction and selection are usually the first steps (Hu et al., 2016; Hojjati, S. H. et al., 2017; Westman, E. et al., 2012; Young, J. et al., 2013; Ye, J. et al., 2012). Most recently, deep learning is introduced to AD research. Deep Neural Network (DNN) model has been successfully implemented in the broad computer vision domains for decades (LeCun, Y. et al., 1998; LeCun, Y. et al., 2015). Related to AD, most efforts are to take the deep learning model as a feature extractor where generic (low-level) and/or problem-specific (high-level) features are extracted from layer to layer. The earlier layers of a deep model contain more generic features that could be used for many domains and the features from later layers are more domain-specific (Nogueira, K. et al., 2017). The features are used in different machine learning models for AD diagnosis (Suk, H. I. et al., 2013; Shi, J. et al., 2017; Suk, H. I. et al. 2014). Other than implementing different machine learning models, researchers further expanded the deep model with one last layer as a classifier for AD diagnosis. For example, Basaia, S. et al., (2019) built a simplified Convolutional Neural Network (CNN) without the need for an activation layer for AD diagnosis. Spasov, S. et al. (2019) designed a parameters-efficient multi-task CNN model for increased generalizability to predict MCI-Converter. Lee, G. et al. (2019) applied a Recurrent Neural Network (RNN), to learn from multi-source data to identify the person with a higher risk of developing AD.

While deep learning opens great opportunities in medical imaging research, its potential is compromised by the limited data available in medicine. Unlike natural images, medical images are rarely available in large quantities. As a result, overfitting is a major obstacle facing the deep learning research community (Srivastava, N. et al., 2014; Lever, J. et al., 2016). One solution to this challenge entails transfer learning (effectively extending knowledge previously learned in one situation to new situations) to implement in deep learning models. In general, deep learning uses transfer learning in the following way: the model is pre-trained on a large labeled dataset (e.g., natural images) to capture the features from images. Then, the model is fine-tuned on the targeted image dataset to extract specific features related to medical images. Therefore, the earliest attempts take a network model as two parts: (1) the first *N* layers are for high-level feature extraction, and (2) the last layer is a classifier. We categorize them as “*N*+1” models. The whole network (*N*+1) is pre-trained on the source domain. In the fine-tuning procedure, the last layer is replaced with the appropriate classification structure tied to the target problem (Hon, M. et al., 2017; Hosseini-Asl, E. et al., 2016). In case they differ greatly, researchers decide to further divide the first *N* layers into (1) first few layers for low-level feature exaction; (2) middle layers for high-level feature extraction. The pre-training is still conducted on the whole network model, fine-tuning on the target domain would involve the middle and last layer of the network (Cheng, D. et al., 2017; Lu, D. et al., 2018). The research reviewed above takes pre-training and fine-tuning as two independent procedures. Lately, researchers start to explore integrating not only the feature extracted from the pre-training but additional features from different sources, into the fine-tuning procedure for improved performance. Liu, T. et al. (2017) fused the features extracted from a pre-trained VGG model with several texture features into a feature pool. Zhang, J. et al. (2017) combined the features extracted from the pre-trained CNN model with handcrafted visual features to classify the type of different medical images (e.g., MRI, CT). Song et al., (2017) generated the Fisher Vector (FV) descriptors integrating the features from the DBN model, the CNN model in an unsupervised manner.

Please note that most existing efforts on transfer learning focus on extracting and transferring features from the pre-training procedure. The outcome (a.k.a. knowledge) from the pre-training process is thus ignored. Here we hypothesize that transferring knowledge from the pre-training to the fine-tuning may benefit the target problem-solving. The knowledge of particular interest in this research is related to a new AD surrogate biomarker (Cole, J. H. et al., 2017). In Cole, J. H. et al., (2017), the researchers trained a deep learning model on MRI neuroimages from cognitively normal subjects to predict each subject’s biological age (B-Age). The trained model was then used to predict the B-Ages for the subjects with brain disease. Under the assumption that B-Age shall align well with chronological age (C-Age) for healthy subjects and the B-Age and C-Age for individuals with brain diseases shall present notable differences, the difference (termed Δ_age_) was used to detect group differences between diseased cohort vs. cognitively normal cohort (Cole, J. H. et al., 2017). Motivated by this initial success from Δ_age_, we are interested in exploring the predictive power of this surrogate marker in classifying MCI converter vs. non-converter on an individual base. Specifically, we propose a new deep learning model named Age-adjust neural network (AD-NET). In the AD-NET, we revisit the transfer learning and propose dual purposes from the pre-trained model: (1) feature transferring: similar to existing research from literature, the pre-trained model without the last layer is used as feature extractor; (2) knowledge transferring: the whole pre-trained model is kept into the fine-tuning stage to transfer the knowledge captured in the age prediction process. Instead of simply appending the Δ_age_ as an additional feature to the CNN model, we propose a composite parameter taking into account the effects from both the group and the individual level to adjust the prediction. Experiments are conducted using two public neuroimaging datasets (IXI (IXI Dataset) and ADNI (F. Li et al., 2018)). We compare our proposed AD-NET with eight existing methods, including Logistic Regression, Partial Least Square, Gaussian Process Regression, Support Vector Machine (SVM), and three deep learning models. Our AD-NET achieved the best AUC of 0.81 and comparable accuracy, sensitivity, and specificity, which are 0.76, 0.77, and 0.76, respectively.

## 2 Method

### 2.1 Architecture and pre-training strategy

The schematic illustration of proposed AD-NET architecture is shown in Figure 1. It contains two separate parts: (1) a pre-trained network for feature extraction and age prediction; and (2) a fine-tuned network to transfer both features and knowledge from age prediction for MCI converter prediction.

**Figure 1.**
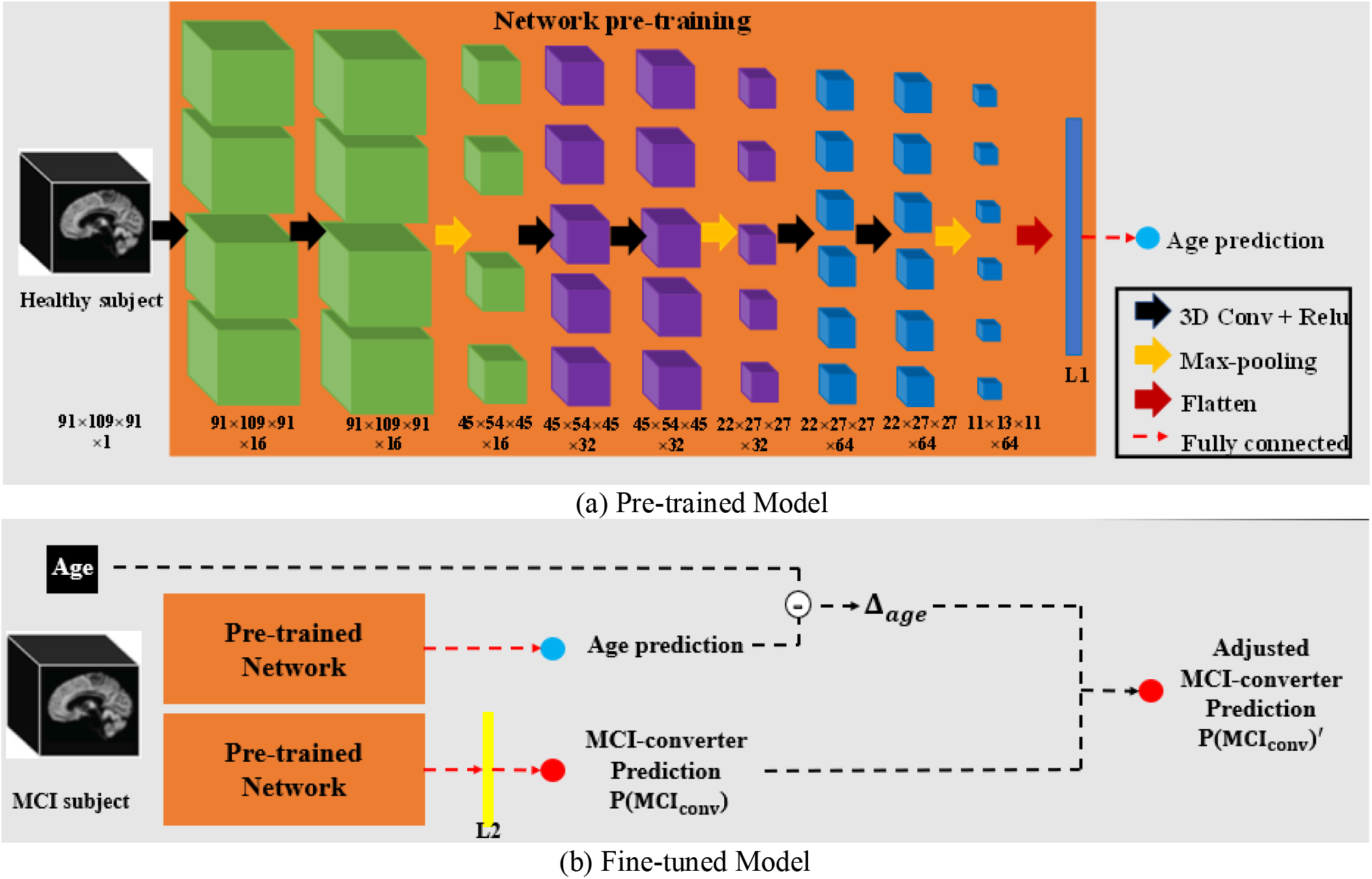
The architecture of the proposed AD-Net. 3D boxes represent input and feature maps. The arrows represent network operations: the black arrow indicates 3D convolutional operation followed by a rectified linear unit (ReLU) activation function; orange arrow represents max-pooling operations; red arrow represents the flatten operation; dotted red arrow represents fully connected layers; purple square represents the regression outputs for predicted B-Age; blue square represents classification outputs for MCI-Converter probability; layers within dotted square form a building block, and there are 3 repeating blocks (block×3) for feature extraction before flatten layer.

**Figure 1.**
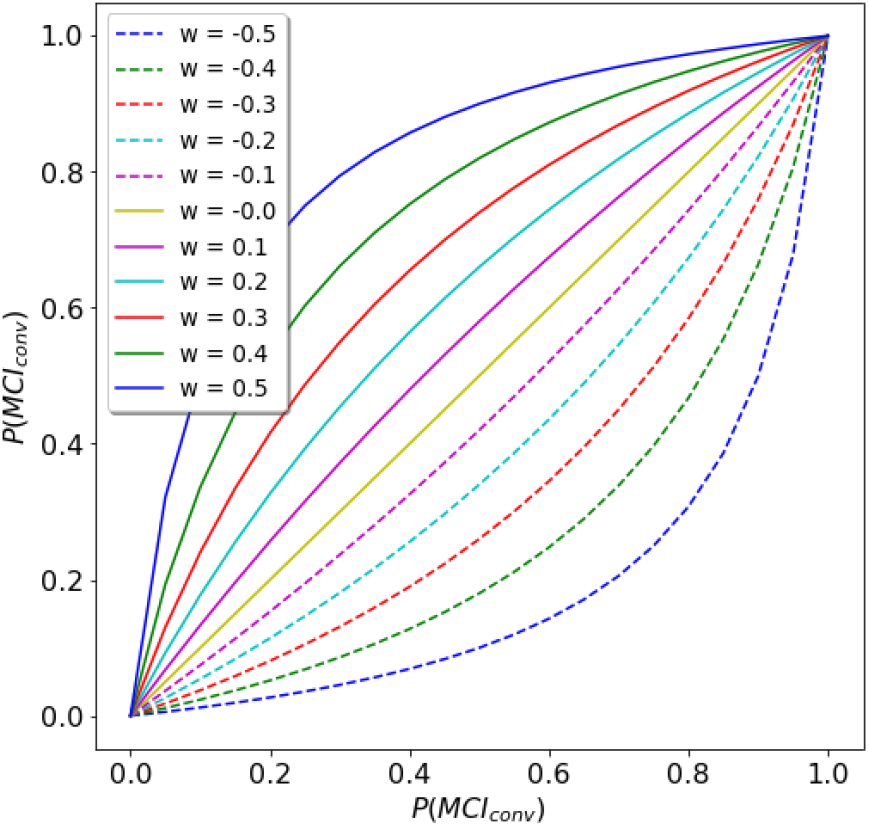
Curves for *P*(*MCI*_*conv*_)′ vs. *P*(*MCI*_*conv*_) under different settings of *w*^*i*^ (*r* =0.8)

Figure 1a is the pre-trained network. It takes 3D MR images from cognitively normal subjects as inputs and predicts age and extracts related features. The size of the input 3D MRI is 91×109×91. It contains repeated 3 blocks, within each block, there are two (3×3×3) convolutional layers and one max-pooling layer; each convolutional layer is followed by a rectified linear unit (ReLU) layer. The number of feature channels is set to be sixteen for the first block and is doubled for each subsequent block. The output of the last block is flattened into one dimension (layer L1 colored with blue in Figure 1). To clarify, within this context, the “flatten” means the vectorization of the multi-dimensional matrix in the last block. This layer is fully connected to one single output with a linear activation function. A dropout layer with a rate equals to 0.2 (as in (He, K. et al., 2016; Gao, F. et al., 2019)) is added to avoid potential overfitting.

The architecture of the fine-tuned model is shown in Figure 1b. Specifically, the L1 layer is fully connected (with dropout rate = 0.2) to the L2 layer, which is connected (with dropout rate = 0.2) to the final single output with a sigmoid activation function for MCI conversion prediction. L2 layer is added to make feature transformation from age prediction task to produce the initial output of MCI-Converter prediction task (*P*(*MCI*_*conv*_)). To serve the knowledge transfer purpose, the whole pre-trained model is kept (including L1) to predict the B-Age, Δ_age_ is then derived and used to adjust MCI prediction *P*′(*MCI*_*conv*_).

For the AD-Net, in the pre-training procedure, the parameters within 3D blocks, layer L1 and B-Age prediction are trained through the age prediction task. In this procedure, a dataset of 900 3D MRI images from cognitively normal subjects was used. In the fine-tuning procedure, the parameters within the pre-trained network were kept fixed, 200 MRI 3D images from MCI patients were used to tune only parameters within the L2 layer to transfer features learned by age prediction task for the MCI-converter prediction task with.

### 2.2 Aging adjustment in fine-tuning procedure

For subject *i*, the chronological age (C-Age) 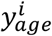 is known as a prior. Given one output from the AD-NET pre-trained model being biological age (B-Age) prediction, that is, 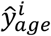, we define 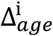 as:

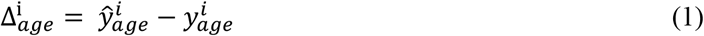

Next, with the 3D neuroimage, AD-NET outputs the risk of the patient *i* to be an MCI-converter 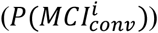 or a non-converter 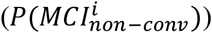, we have 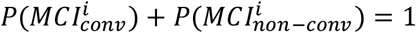.

Under the hypothesis that Δ_*age*_ is strongly correlated to the risk of developing brain disease (Cole, J. H. et al., 2017), most MCI subjects’ brain appears older than that of healthy subjects. Also, MCI Converters’ more likely have a larger Δ_*age*_ than that of MCI non-converter subjects, so Δ_*age*_ represents the risk of developing a brain disease. To utilize this idea, we propose to adjust the probability of an MCI subject *i* converting to 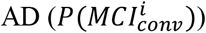 with 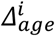. The basic idea is, for patient *i*, if the predicted B-Age is greater than its C-Age, that is, 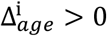, this individual has an increased risk of converting to AD, and vice versa. To model this idea, let take 0.5 as the midpoint (e.g., 50% risk of conversion), the conversion risk with respect to the non-conversation risk, in the form of a ratio, is adjusted as:

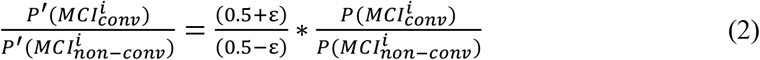

where, 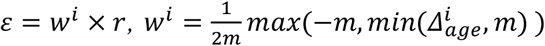, *m* is pre-defined normalizer to remove outlier impact (e.g., extreme large Δ_*age*_), *r* is the correlation between all Δ_*age*_ and MCI-Converter labels, and 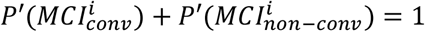. Here *ε* is a composite parameter of two scalars: a global scalar *r* and a subject-dependent scalar *w*^*i*^. Global scalar *r* (*r* ∈ [-1, 1]) is derived as the Pearson correlation between the Δ_*age*_ with the patient’s status. A total positive linear correlation exists for *r* being 1, and total negative linear correlations for *r* being -1, no correlation for *r* being 0. In this study, we would expect to have *r* being positive value to describe the general relationship between the Δ_*age*_ and the patient status on the group bases. Scalar *w*^*i*^ is to measure the normalized deviation level of 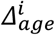 for subject *i. w*^*i*^ is proportional to 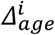, and it is normalized to the range of -0.5 to 0.5 by a pre-defined normalizer *m*.

To better illustrate the effects of *w*^*i*^ and *r* in adjusting 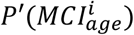, we plot *P* (MCI_conv_) vs. *P*′(MCI_conv_) under different settings of *w*^*i*^ with a given *r* (see Figure 2 with *r=*0.8). Here we only discuss the scenario where *r* is positive given the cohorts being interested are MCI subjects (same holds true when *r* is negative), and *w*^*i*^ can be both negative and positive. From Figure 2, we observe two properties:

**Figure 2.**
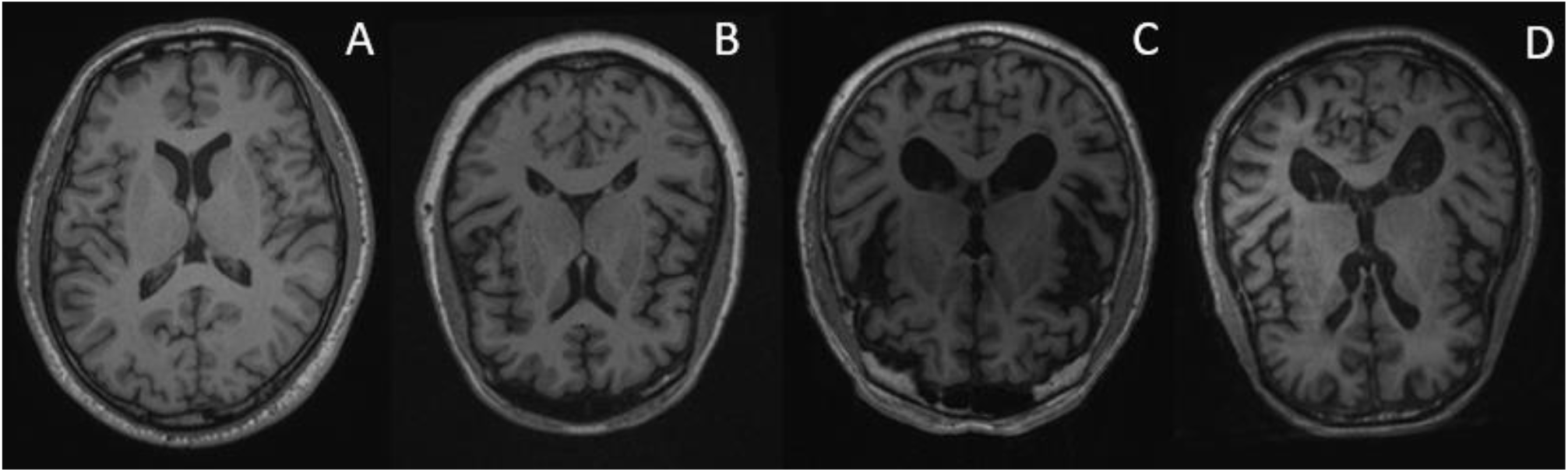
Sample slices from input T1-weighted MRI imaging after the minimal pre-processing procedure. A) cognitively normal subject from the IXI dataset. B) cognitively normal subject from the ADNI dataset. C) MCI Non-Converter subject from the ADNI dataset. D) MCI-Converter subject from the ADNI dataset.

1. For a positive *w*^*i*^ (an individual with increased risk of conversion), *P*′(MCI_conv_) increases as *w*^*i*^ increases. That is, the larger the *w*^*i*^ is, the greater adjustment made from *P*(*MCI*_*conv*_) to *P*′(MCI_conv_). For negative *w*^*i*^, *P*′(*MCI*_*conv*_) decreases as *w*^*i*^ decreases. That is, the smaller the *w*^*i*^ is, the greater adjustment made from *P*(*MCI*_*conv*_) to *P*′(MCI_conv_). This is consistent with our hypothesis, that is, given *w*^*i*^ is proportional to 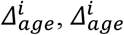 thus *w*^*i*^ is positively correlated with the AD conversion risk.
2. The adjustment has more effects for subjects with *P*(MCI_conv_) falling in the middle of the distribution (e.g., 0.4 - 0.6) than that at the two sides (e.g., 0-0.1 and 0.9-1.0). We believe this is a desirable property indicating the adjustments can strengthen the differentiation power for the subjects who were not certain on determining the conversion risks.

In the fine-tuning model, the age-related information from the pre-training is transferred. Together with the features from the pre-training model, the risk of the subject converting to AD is predicted. A comprehensive comparison experiment is conducted and is discussed in the next sub-sections.

### 2.3 Data

#### 2.3.1 Dataset I for age prediction

All neuroimaging data used in the study are T1-weighted MRI. The dataset used in the pre-training procedure for age prediction task included 847 cognitively normal subjects. The datasets were obtained from two sources, and we conducted the pre-processing procedure to ensure consistency among images from different cohorts. The first data source was Alzheimer’s Disease Neuroimaging Initiative (ADNI) dataset (F. Li et al., 2018), the ages range from 56-89. The ADNI is launched aiming at finding the relationship between progression of mild cognitive impairment (MCI) to early Alzheimer’s disease (AD) with biomarkers, from MRI, PET or clinical and neuropsychological assessments. ADNI enrolled a large cohort of participants (Weiner, M. W. et al., 2015) with the majority of participants having PET, MRI as well as clinical information (including age) available. Data were collected as a longitudinal multisite study by the National Institute of Biomedical Imaging and Bioengineering, the National Institute on Aging, the Food and Drug Administration, non-profit organizations, and private pharmaceutical companies. Approved consent was obtained from all patients, and the ADNI has been approved by the institutional review board at all sites. We selected 256 subjects who were cognitively normal (CDR=0) and amyloid negative (based on florbetapir PET) at the time of MRI scan from ADNI for the pre-training. In order to increase the size of the training dataset and widen the age range for robust age prediction, we obtained 581 cognitively normal subjects from a second data source: Information eXtraction from Images (IXI) public dataset (IXI Dataset). The subjects from the IXI dataset were obtained from 3 different hospitals in London: Hammersmith Hospital, Guy’s Hospital and Institute of Psychiatry. Approved consent was obtained from all patients, and IXI was approved by an ethics committee at all sites. For each subject, personal information such as sex, height, weight, occupation and age were included.

### 2.3.2 Dataset II for MCI-conversion prediction

The dataset used in the fine-tuning procedure for the MCI conversion prediction task was obtained from ADNI. All subjects have the status as being either converter or non-converter. The dataset included a total of 297 subjects. These subjects were diagnosed as MCI during the baseline visit. Among the 297 subjects, 168 were MCI-converters, and 129 subjects were MCI non-converter. The MCI-converter and MCI non-converter subjects were labeled through the following logic: a subject was labeled as MCI-converter if the subject was diagnosed as MCI and converted to AD during a three-year follow-up, and a subject was labeled as MCI non-converter if the subject was diagnosed as MCI at both baseline and the three-year follow-up. Those subjects whose diagnosis was missing at the three-year follow-up were excluded.

### 2.4 Pre-processing

We used MRIcron (https://www.nitrc.org/projects/mricron) to convert DICOM files to NIfTI format and conducted rigid registration to MNI152 (Fonov, V. S. et al. 2009) space to ensure consistency of position and orientation. The images were resampled using cubic spline interpolation, to transfer data acquired from different studies into the same voxel sizes and dimensions (1mm3, 182×218×182). Examples of the different data used in the study are shown in Figure 2.

### 2.5 Design of Experiments

#### 2.5.1 Experiment I: Pre-training and Age Prediction Task

The objective of this experiment I is to develop a pre-training model for age prediction. In this experiment, 84 (10%) subjects were randomly selected from Dataset I as a blind testing dataset, the remaining 763 subjects were used as the training dataset. The proposed AD-NET was trained using mean squared error (MSE) as loss function, Adam (Kingma, D. P. et al., 2014) was used as the optimizer to solve the problem. We set the hyper-parameter as: (1) learning rate was 0.01; (2) learning rate decay equaled to 0.005; (3) training batch was 16; and (4) training iteration was 200.

#### 2.5.2 Experiment II

We conduct the second experiment using the surrogate marker to predict MCI conversation risk. 5-fold cross-validation was conducted to evaluate AD-NET’s performance on the MCI-Converter prediction problem. The parameters of the deep network obtained from the pre-training procedure were kept the same for the age prediction. The parameters of L2 layer were fine-tuned to convert the age prediction features for the MCI converter prediction problem. The AD-NET was fine-tuned using cross-entropy as loss function and Adam optimizer (Kingma, D. P. et al., 2014). Other parameters are selected based on the best performance: (1) learning rate was 0.01; (2) learning rate decay equaled to 0.005; (3) training batch was 16; and (4) training iteration was 50. The area under a receiver operating characteristic curve (AUC), accuracy (ACC.), sensitivity (SEN.), and specificity (SPE.) were calculated to measure the prediction power of our model from different aspects.

For comparison purpose, we implemented two competing methods, which were pre-trained through the same procedure as AD-NET: Transfer learning CNN model (TL-CNN) and Transfer learning CNN model with Δ_age_ as additional features (TL-CNN-Δ_age_). The architecture of TL-CNN is the same as our-proposed AD-NET, the major difference is that during the fine-tuning procedure, neither C-Age information nor predicted B-Age from pre-training procedure was included. This deep learning architecture is well-studied in a number of medical image applications such as age prediction (Cole, J. H. et al., 2017), breast cancer classification (Gao, F., et al. 2018) and medical imaging synthesis (Li, R., et al., 2014). In TL-CNN-Δ_age_, the Δ_age_ for each subject was calculated after the pre-training procedure. During the fine-tuning procedure, Δ_age_ was simply added as an additional input into the last layer (layer L2 in Figure 1). In addition, six existing methods, which used the same ADNI dataset, from the literature were chosen for comparison. These included both traditional machine learning models (e.g., logistic regression and SVM) and deep learning models. Also, we further explore the role of Δ_age_ in the MCI-conversion prediction problem among different age groups.

## 3 Results

### 3.1 Experiment I: Pre-training and Age Prediction Task

In experiment I, the proposed model achieved MSE of 187.16 and Mean Absolute Error (MAE) of 11.17 on the training dataset. The Pearson Correlation (*PC*) between C-Age (*y*_*age*_) and predicted B-Age (*ŷ*_*age*_) was 0.75. On the testing dataset, we had MSE=196.42, MAE=12.28, *PC*=0.67. For illustration purposes, we include the plot of C-Age vs. predicted B-Age for both training dataset (Figure 3A) and testing dataset (Figure 3B). We conclude that after pre-training, the AD-NET for age prediction can successfully capture the correlation between raw MRI image and C-Age among the healthy subjects as demonstrated by the Pearson correlation and the following figure.

**Figure 3.**
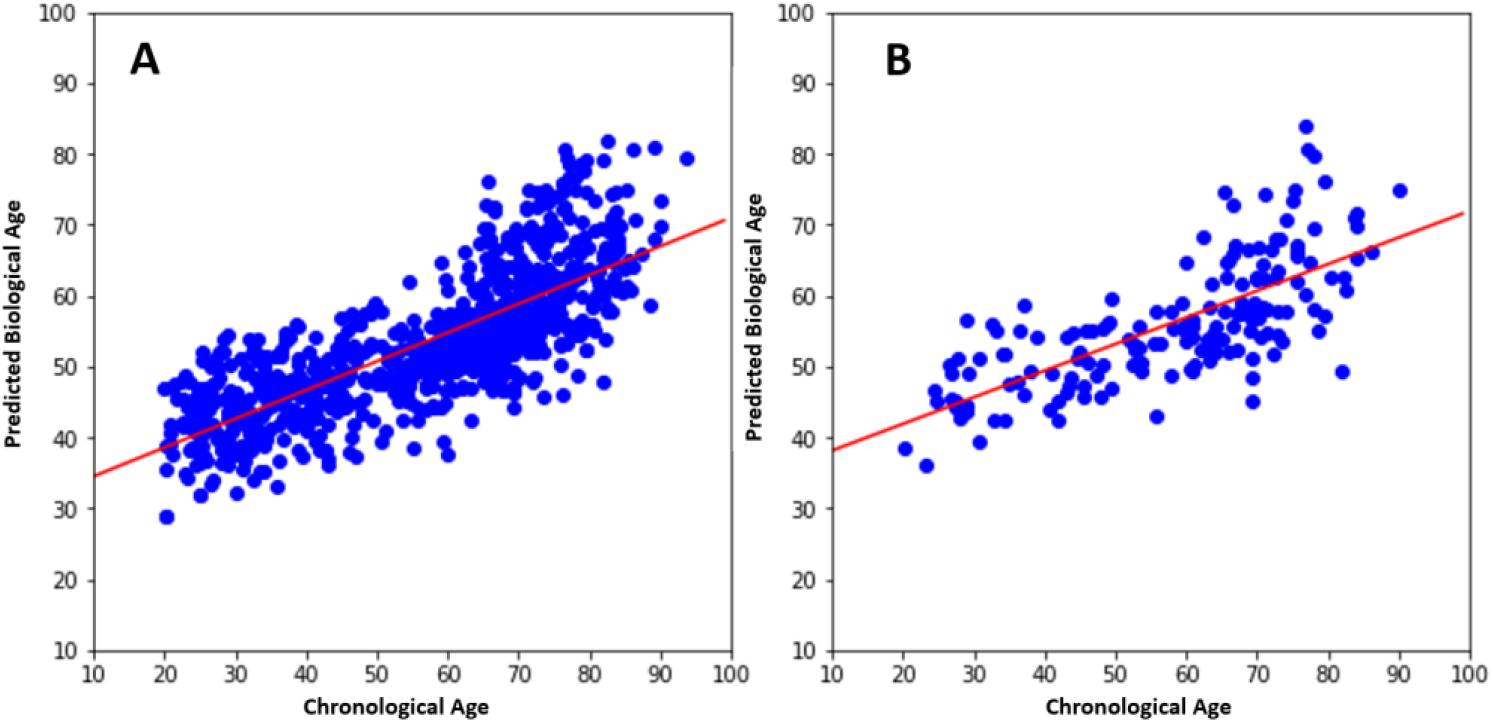
Plot of chronological age (C-Age) vs. predicted biological age (B-Age): A) training dataset (MSE=187.16, MAE=11.17, PC=0.75), B) testing dataset (MSE=196.62, MAE=12.28, PC-0.67). Red lines are the fitted linear regression, respectively.

We do recognize our model on aging prediction may not be as optimum as that from (Cole, J. H. et al., 2017). We also observe the predicted B-Age tends to be younger than the C-Age. There is certainly room for model improvement. With the consistent performance of the model on the training vs. blind testing dataset, one conclusion we can draw is the model is robust. Given the focus of this study is to demonstrate the advantages of surrogate biomarkers from age for MCI converter prediction, we decide to leave the age prediction model improvement as a future research effort.

Now we fed all subjects in Dataset II into the pre-trained model and obtained predicted age for each MCI subject. The *Δ*_*age*_ for each subject was derived using equation (2). We conducted a t-test between the MCI-converters and MCI non-converters at different age groups (Table 2). From this group-based study, it is interesting to observe that there is statistical significance between the MCI converters vs. MCI non-converters under the age group ranges from 55-90, 60-90, 65-90 and even from 70-90. For the cohorts aging 75+, the statistical difference diminishes. We contend the power of this age surrogate biomarker may be weakened in the older senior population. To get the composite parameter *ε* for equation (2), we need to derive the Pearson Correlation *r*. As discussed above, our model tends to predict younger B-Age comparing to C-Age, the mean *Δ*_*age*_ for all subjects in Dataset II is -16.64. Ideally, we would like the mean *Δ*_*age*_ close to zero, to utilize positive or negative *Δ*_*age*_ (thus the *w*^i^) to adjust *P*(*MCI*_*conv*_). Therefore, we shifted the *Δ*_*age*_ with -16.64. Figure 5 shows the distribution of *Δ*_*age*_ after this adjustment. In order to avoid impacts from the extreme values of adjusted *Δ*_*age*_, z-score method is used to identify potential outliers (Ben-Gal, I. 2005). Any samples outside the ±2 standard deviation of *Δ*_*age*_ range were adjusted to these threshold values to keep model training less sensitive to those extreme values (i.e., m=17, that is, adjusted *Δ*_*age*_will be within ±17 in our setting). Given the Pearson correlation (*PC*) between *Δ*_*age*_ and MCI-Converter labels is 0.15, we set *r* = 0.15.

**Table 1.** Demographic information for subjects in Dataset I and Dataset II

|  | Dataset I | Dataset II |
| --- | --- | --- |
| Data Source | ADNI and IXI | ADNI |
| Sample Size | 847 | 292 |
| Sex (M/F) | 395/452 | 185/107 |
| Age Mean ± std (years) | 56.86 ± 18.34 | 74.84 ± 7.24 |
| (Range in years) | (18 – 94) | (55 – 89) |
| Apolipoprotein e4<br>(NC/HT/HM) | N/A | 129/128/35 |
| Education Mean ± std<br>(Years)<br>(Range) | N/A | 16 ± 3<br>(7 – 20) |
| MMSE Mean ± std<br>(Range) | N/A | 27.04 ± 1.76<br>(23 – 30) |

NC = Non-carrier; HT = Heterozygote; HM = Homozygote; N/A = Not available.

**Table 2.**
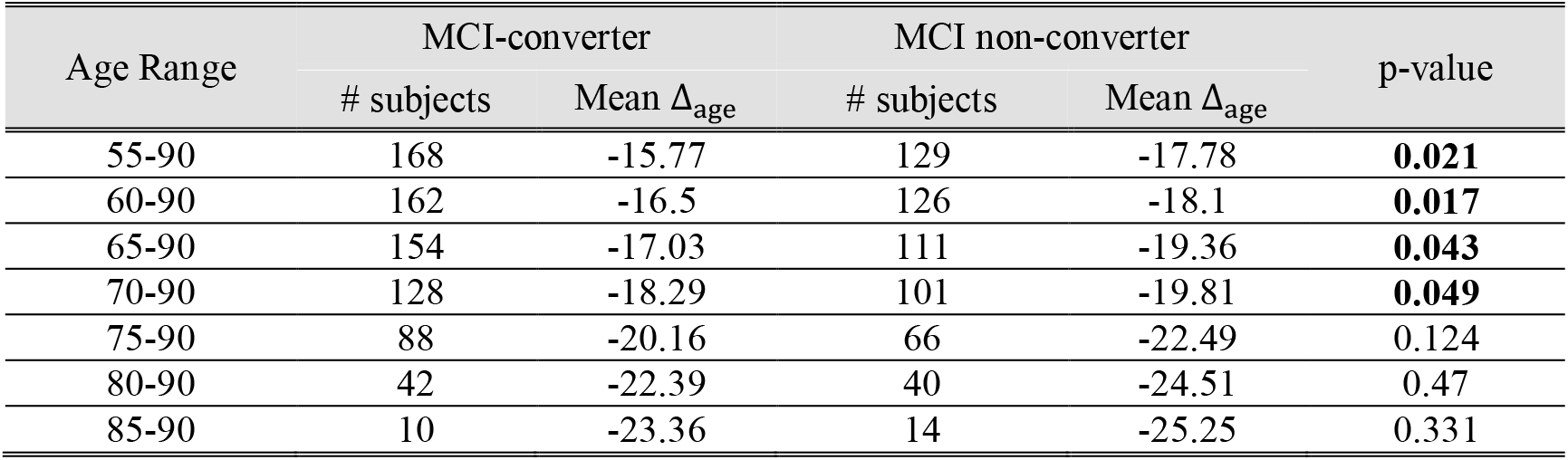
P-values of t-test on age-gap between MCI-converter vs. non-converter group at the different age range

**Figure 5.**
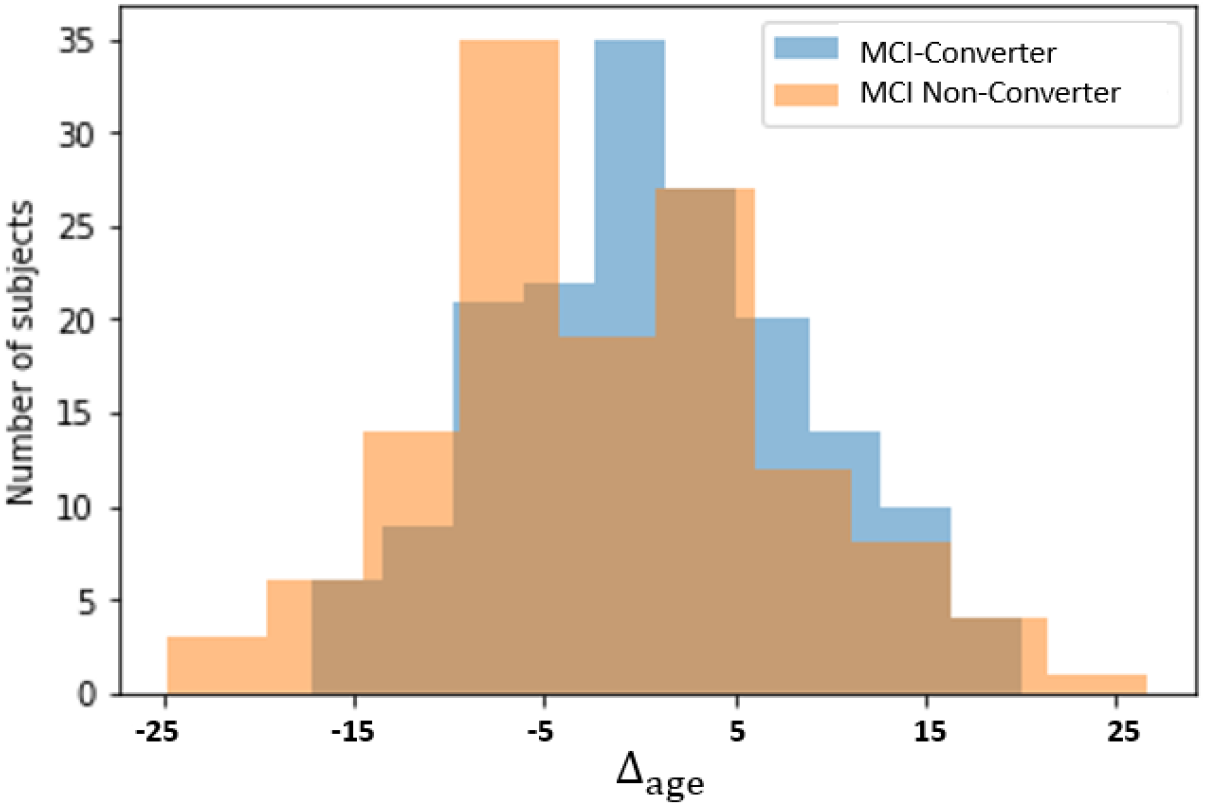
Distribution normalized *Δ*_*age*_ values for MCI-Converter and MCI Non-Converter groups

In this first experiment, the accuracy of AD-NET in age prediction task and the potential of biomarker Δ_age_ in differentiating MCI-converter vs. MCI non-converter on the group bases are validated.

### 3.2 Experiment II: MCI-Converter Prediction Task

In this experiment, we incorporated *Δ*_*age*_ as a surrogate biomarker to predict MCI conversion risk of each subject. Table 3 summarizes the performance comparison among the models from the literature and our proposed model. Four measures, AUC, Accuracy, Sensitivity, and Specificity are used.

**Table 3.** Comparison Results on AUC, Accuracy, Sensitivity, and Specificity

| METHODS | MODEL | DATA | AUC | ACC. | SEN. | SPE. |
| --- | --- | --- | --- | --- | --- | --- |
| Logistic/Cox regression (Ewers, M. et al. 2012) | ML | Structural MRI+CSF+ Neuropsychological testing | NA | 0.77 | 0.82 | 0.73 |
| Orthogonal partial least squares (Westman, E. et al., 2012) | ML | Structural MRI + CSF | 0.76 | 0.69 | 0.74 | 0.63 |
| Gaussian Process (Young, J. et al., 2013) | ML | Structural MRI + CSF + PET + APOE | 0.80 | 0.74 | 0.79 | 0.66 |
| SVM (Liu, F., et al., 2014) | ML | Structural MRI + PET | 0.70 | 0.68 | 0.65 | 0.70 |
| SAE + Logistic regression (Liu, S. et al., 2014) | DL/ML | Structural MRI + PET | NA* | 0.54 | 0.52 | 0.87 |
| Deep polynomial network + SVM (Shi, J. et al., 2017) | DL/ML | Structural MRI + PET | 0.80 | 0.79 | 0.68 | 0.87 |
| TL-CNN (Cole, J. H. et al., 2017) | DL | Structural MRI | 0.76 | 0.73 | 0.68 | 0.77 |
| TL-CNN-$\Delta_{age}$ | DL | Structural MRI + Age | 0.77 | 0.77 | 0.80 | 0.73 |
| AD-NET | DL | Structural MRI + Age | 0.81 | 0.76 | 0.77 | 0.76 |

\*NA: not reported in the literature

We have four observations from Table 3. First, traditional machine learning models (e.g., Westman, E. et al., 2012; Young, J. et al., 2013; Ewers, M. et al. 2012; Liu, F., et al., 2014) require additional information (e.g., clinical testing scores, APOE) to achieve comparable performance as deep learning models on imaging data only. We thus conclude this demonstrates the advantage of deep learning models, which may better explore the power of neuroimaging for disease diagnosis. Second, in handling neuroimaging data alone, (Liu, S. et al., 2014; Shi, J. et al., 2017) are the combination of a deep model with a machine learning model where deep learning models are feature extractors. While both have good specificity (SPE.), the performance on sensitivity (SEN.) is less than desirable. Here we want to emphasize that sensitivity is of more clinical importance in this study as the goal is to identify the MCI converters early for effective interventions. In addition, (Liu, S. et al., 2014; Shi, J. et al., 2017) would require both MRI and PET for such performance. In using MRI imaging alone, we compare TL-CNN (Cole, J. H. et al., 2017), TL-CNN-Δ_age_ with our proposed AD-NET. Our third observation is with Δ_age_ added, TL-CNN-Δ_age_ and AD-NET outperforms TL-CNN in terms of overall performance metrics (ACC. and AUC). This demonstrates the potential of Δ_age_ as a surrogate marker for the MCI conversion prediction problem. In comparing AD-NET vs. TL-CNN-Δ_age_, we have 0.76 (ACC.), 0.77 (SEN.), 0.76 (SPE.) for AD-NET and 0.77 (ACC.), 0.80 (SEN.), 0.73 (SPE.) for TL-CNN-Δ_age_. Though it seems TL-CNN-Δ_age_ outperforms AD-NET on sensitivity (0.80 vs. 0.77), t-test shows there exists no statistically significant difference (*p* = 0.24). For AUC, t-test indicates AD-NET significantly outperforms TL-CNN-Δ_age_(*p* = 0.02). We conclude AD-NET achieves comparable performances in terms of accuracy (ACC.), sensitivity (SEN.) and specificity (SPE.) as that from TL-CNN-Δ_age_ and outperformance TL-CNN-Δ_age_ in AUC indicating the robustness of AD-NET.

We want to emphasize that in medical research, AUC is considered as a more consistent metric with better discriminatory power comparing to accuracy (Lu, D. et al., 2018). As seen in Table 3, AD-NET has the highest AUC among all the models.

Table 4 depicts the performance of MCI-conversion prediction among different age groups and how Δ_age_ play a role. As observed, for the younger group (55-75), the AUC values are 0.74, 0.75 and 0.79 for TL-CNN, TL-CNN-Δ_age_ and AD-NET, respectively. For the older senior group (75-90), group-based study (Table 2) indicates the diminished power of *Δ*_*age*_ as a surrogate age marker to distinguish MCI converter vs. non-converter. All three deep learning models show the promises in predicting the conversion risk of each individual subject. In addition, AD-NET consistently outperforms TL-CNN, TL-CNN-Δ_age_ for all subjects (see Table 3) and the separated age group (see Table 4). We conclude AD-NET, a deep learning model enabled by knowledge transfer has the potential for AD early detection.

**Table 4.** AUC performance for TL-CNN, TL-CNN-*Δ*_*age*_, AD-NET under for different age group

| AGE RANGE | # OF SUBJECT | METHODS | AUC |
| --- | --- | --- | --- |
| 55-75 | 80 MCI converters | TL-CNN (Cole, J. H. et al., 2017) | 0.74 |
| | vs. | TL-CNN-$\Delta_{age}$ | 0.75 |
|  | 63 MCI non-converters | AD-NET | 0.79 |
| 75-90 | 88 MCI converters | TL-CNN (Cole, J. H. et al., 2017) | 0.80 |
| | vs. | TL-CNN-$\Delta_{age}$ | 0.80 |
|  | 66 MCI non-converters | AD-NET | 0.83 |

## 4 Discussion

While AD-NET shows the potentials in MCI conversion risk assessment via the surrogate age biomarker, it has limitations. First, as discussed in the first experiment, we do recognize our deep learning model on aging prediction may underperform the model from Cole, J. H. et al., 2017. We contend this is because the focus of this study is to demonstrate the power of the surrogate age biomarker. This leads to our imminent future work on improving the performance of the age prediction. One solution is to gather more neuroimages and dedicate efforts to model tuning. We believe with improved performance on age prediction, MCI-conversion risk assessments will potentially be improved. The second limitation is that the AD-NET in this research focuses on neuroimaging data only. We plan to incorporate information such as sex, gene, clinical test scores, etc. from each subject as the competing methods did, into the model to further improve the performance. The third limitation is related to global scalar r, which is pre-defined based on prior knowledge. We plan to identify the optimal r during the model training process to explore the impact of the global scalar on the model performance.”

In this research, we propose a new deep learning model: AD-NET (Age-adjust neural network). One contribution of AD-NET is to transfer the knowledge captured in the pre-training, specifically, a surrogate biomarker Δ_age_ (the difference between chronological age and predicted biological age). The knowledge-based transfer learning not only saves training resources but also improves prediction accuracy. Our second contribution lies in a novel age adjust procedure. Instead of simply adding Δ_age_ as an additional feature to the deep model, we introduce a composite parameter, *ε* = *w*^*i*^ × *r*, considering the effects of both the group (*r*) and the individual subject (*w*^*i*^) to adjust the prediction (see equation (2)). Subject-dependent scalar, *w*^*i*^, is smoothed 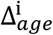 by removing outlier impact. It is straightforward to have *w*^*i*^ included in the prediction model. Here we provide more discussions on our investigation on the *r*.

Global scalar *r* is the Pearson correlation between the Δ_*age*_ with the patient status (MCI converter vs. MCI non-converter) for the cohort, *r* ∈ [-1, 1], *r* = 1 for a total positive linear correlation, *r* = -1 for a total negative linear correlation, *r* = 0 for no correlation. We conducted experiments for sensitivity analysis of *r* for the prediction (see Figure 6). When *r* = 0, we have *ε* = 0, AD-NET essentially is the same as TL-CNN, we have AUC=0.76. When *r* = 1, we have *ε* = *w*^*i*^, AUC has the lowest value as 0.69. Note AD-NET under *ε* = *w*^*i*^ differs from TL-CNN-Δ_age_ due to the use of equation (2) in the prediction adjustment. It is interesting to observe the best AUC is obtained when *r* ranges [0.1, 0.3]. The Pearson correlation *r*, in this study, 0.15 is one of the best scenarios. We contend this may be because the prediction deep learning model is cross-trained on the subject group. The model parameters are set capturing the underlying correlations to the subject group. The empirical experiment demonstrates that incorporating a Pearson correlation to the composite parameter helps improve the prediction performance. It is our intention to conduct theoretical research as immediate next step to understand the interrelationship among Pearson correlation *r*, subject-dependent scalar, *w*^*i*^, composite parameter *ε* and prediction performance.

**Figure 6.**
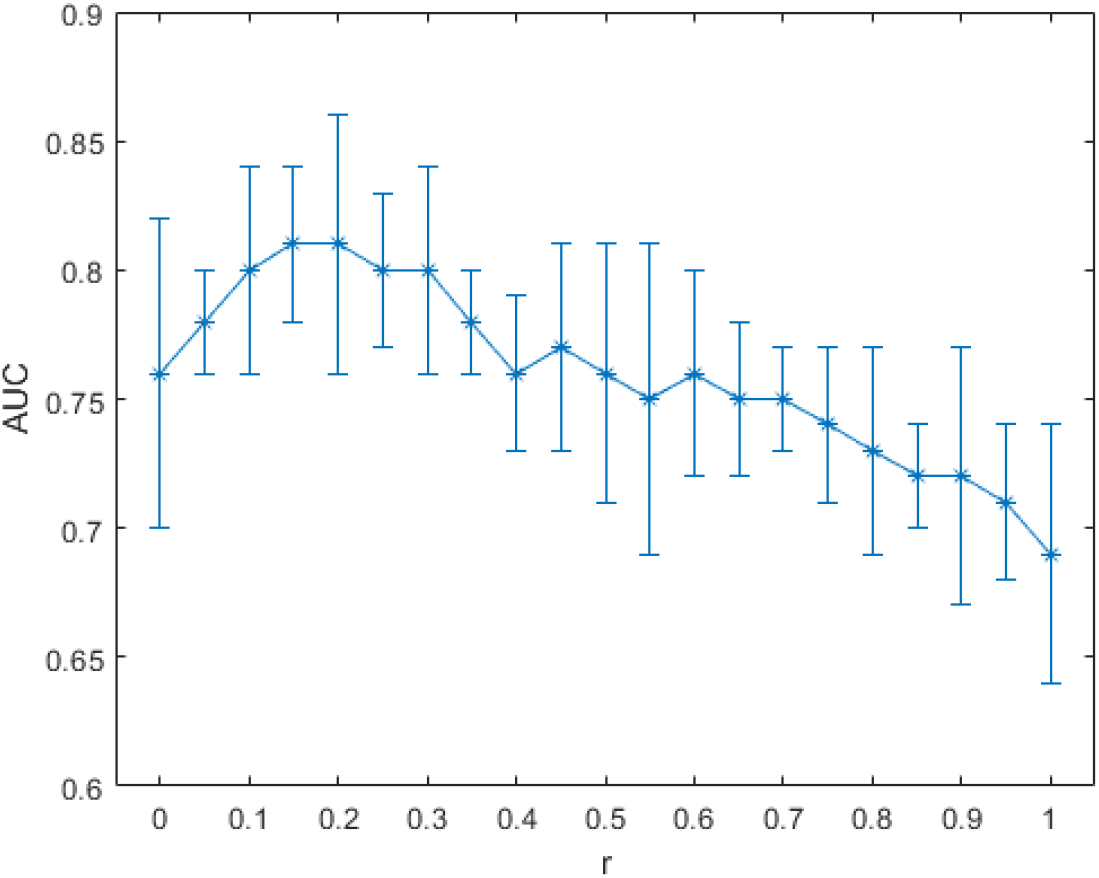
Sensitivity Experiments on global scalar *r* on AUC values for MCI converter vs. non-converter prediction

## 5 Conclusions

Our proposed AD-NET extends traditional feature-based transfer learning with knowledge transfer capability. Surrogate biomarker Δ_age_, captured through pre-training, is adjusted by global and individual factors for the fine-tuning stage. We compare AD-NET with 8 classification models from literature using the same public neuroimaging dataset. Experimental results show that the proposed AD-NET outperforms all the competing models on AUC in predicting the MCI converter vs. non-converter. While promising, there is room for improvement. Other than conducting theoretical analysis on the Pearson correlation as discussed above, we plan to improve the pre-training network for age prediction. We expect the improved age prediction will help improve the MCI conversion predictions further which may facilitate future clinical trial design and improved management of the disease. Another direction for future work is to explore the model performance on multiple neuroimaging modalities, e.g., both MRI and PET.

## Data Availability

Data used in the preparation of this article were obtained from the Alzheimer’s Disease Neuroimaging Initiative (ADNI) database (adni.loni.usc.edu). As such, the investigators within the ADNI contributed to the design and implementation of ADNI and/or provided data but did not participate in analysis or writing of this report. A complete listing of ADNI investigators can be found at: http://adni.loni.usc.edu/wp-content/uploads/how_to_apply/ADNI_Acknowledgement_List.pdf
Data used in preparation of this article were also obtained from the Infomation eXtraction from Images (IXI) public dataset (IXI dataset), which was obtained from three different hospitals in London: Hammersmith Hospital, Guy's Hospital and Institute of Psychiatry (https://brain-development.org/ixi-dataset/). As such, the investigators associated with IXI contributed to the design and implementation of IXI dataset and/or provided data but did not participate in analysis or writing of this report.
The relevant ethical guidelines have been followed. All necessary IRB and/or ethics committee approvals were obtained, and details of the IRB/oversight body have been included in the manuscript.

## Acknowledgement

This work is supported in part by R01AG031581, P30AG019610, DHS and the State of Arizona. ADHS Grant No. CTR040636 (previously ADHS Grant No. ADHS14-052688). Dr. Su is also funded by BrightFocus Foundation ADR A2017272S, Alzheimer’s Association AARG-17-532945. The funding sources did not play a role in study design, the collection, analysis and interpretation of data, writing of the report; or in the decision to submit the article for publication.

Data collection and sharing for this project was funded by the Alzheimer’s Disease Neuroimaging Initiative (ADNI) (National Institutes of Health Grant U01 AG024904) and DOD ADNI (Department of Defense award number W81XWH-12-2-0012). ADNI is funded by the National Institute on Aging, the National Institute of Biomedical Imaging and Bioengineering, and through generous contributions from the following: AbbVie, Alzheimer’s Association; Alzheimer’s Drug Discovery Foundation; Araclon Biotech; BioClinica, Inc.; Biogen; Bristol-Myers Squibb Company; CereSpir, Inc.; Cogstate; Eisai Inc.; Elan Pharmaceuticals, Inc.; Eli Lilly and Company; EuroImmun; F. Hoffmann-La Roche Ltd and its affiliated company Genentech, Inc.; Fujirebio; GE Healthcare; IXICO Ltd.; Janssen Alzheimer Immunotherapy Research & Development, LLC.; Johnson & Johnson Pharmaceutical Research & Development LLC.; Lumosity; Lundbeck; Merck & Co., Inc.; Meso Scale Diagnostics, LLC.; NeuroRx Research; Neurotrack Technologies; Novartis Pharmaceuticals Corporation; Pfizer Inc.; Piramal Imaging; Servier; Takeda Pharmaceutical Company; and Transition Therapeutics. The Canadian Institutes of Health Research is providing funds to support ADNI clinical sites in Canada. Private sector contributions are facilitated by the Foundation for the National Institutes of Health (www.fnih.org). The grantee organization is the Northern California Institute for Research and Education, and the study is coordinated by the Alzheimer’s Therapeutic Research Institute at the University of Southern California. ADNI data are disseminated by the Laboratory for Neuro Imaging at the University of Southern California.

## Author Declarations

The relevant ethical guidelines have been followed. All necessary IRB and/or ethics committee approvals were obtained, and details of the IRB/oversight body have been included in the manuscript

